# Can an online battery match in-person cognitive testing in predicting age-related cortical changes?

**DOI:** 10.1101/2023.04.24.23289014

**Authors:** R. Thienel, L. Borne, C. Faucher, G.A. Robinson, J. Fripp, J. Giorgio, A. Ceslis, K. McAloney, J. Adsett, D. Galligan, N.G. Martin, M. Breakspear, M. K. Lupton

## Abstract

Understanding how cognition and brain structure change across the lifespan is crucial for gaining insight into the healthy ageing process, as well as identifying early signs of neurodegenerative changes. In our recent prospective study of healthy ageing in midlife and older adults^1^, we compared the association of two cognitive batteries with age-related variability in brain morphology. Our findings revealed that online cognitive testing, which is more cost-effective, demonstrated comparable association to sulcal width as comprehensive in person assessment. Both cognitive assays show similarly strong correlations with sulcal width. In addition, it was found that both cognitive assessment assays showed a more pronounced age-related decline in individuals with Aβ burden. These findings suggest that online assessment is able to detect accelerated cognitive ageing comparably to the in-person assay in our preclinical sample, even in the early stages of Aβ accumulation before significant structural brain changes occur. Taken together with their greater cost effectiveness, online cognitive testing could lead to more equitable early detection and intervention for neurodegenerative diseases.

## BACKGROUND

Ageing is associated with substantial changes in cognition^2^ and brain structure^3^. The trajectories of these age-related changes show significant individual differences with major clinical implications^4^ such as early detection of neurodegenerative disorders (e.g., Alzheimer’s Disease). Comprehensive in-person neuropsychological assessments across multiple cognitive domains are routinely used to estimate cognitive status in both research and clinical settings and are considered gold standard. This type of assessment is labour intensive, costly and not always feasible. Accessibility issues are a significant barrier for older adults to receive health care^5^. Recent events, such as the Covid-19 pandemic, have also been found to influence older adults’ access to health care^6^. These considerations raise the importance of reducing barriers to participation in clinical care and research.

Self-administered online cognitive testing offers several advantages over in-person assessments, including greater flexibility, the ability to record accuracy and speed of response with high precision, and better cost-efficiency^7^. The popularity of online neuropsychological tests is rapidly increasing, with the availability of online cognitive batteries having more than doubled in the past decade^8,9^ and large biomedical databases such as the UK biobank (https://www.ukbiobank.ac.uk/) solely relying on computerized testing. Earlier studies initially expressed skepticism about the use of computerised testing, particularly regarding the introduction of environmental confounds and the lack of supervision^10,11^. Nonetheless, research in large samples has shown a strong correlation (Pearson’s r=0.80) between in-person and web-based cognitive testing^12,13^, suggesting potential for high-quality data comparable to in-person testing when quality insurance measures are met. This is excellent considering that test-retest reliabilities of widely used in person neuropsychological tests are highly variable (ranging between r=0.5-0.9 for individual tests, with memory and executive functioning scores often less than r=0.7)^14^. Only few studies have validated the use of online cognitive testing in older adults, but unsupervised web-based tests, including the Stroop task, paired associates learning, and verbal and matrix reasoning, have been shown to yield comparable results to supervised tests administered in a laboratory^15^. Moreover, performance on web-based tests does not appear to be correlated with technology familiarity, an issue previously raised as a potential barrier^15^.

Creyos (previously Cambridge Brain Sciences, CBS), is a widely used online cognitive assessment platform that consists of 12 self-administered tasks, based on well-validated neuropsychological tests adapted for use in a home environment^16^. Creyos is designed to adjust the level of difficulty of each task according to the performance level of the participant, with corresponding increases and decreases in complexity minimising floor and ceiling effects. In addition, data reliability is ensured through ‘validity’ indicators, which flag when the data are outside the expected bounds for the correct and uninterrupted task completion. Creyos has been used in several large-scale epidemiological studies^17,18,19^. There have only been a limited number of studies comparing the use of the Creyos platform with in-person neuropsychological testing in older individuals (aged ≥40years), and these have used small sample sizes and non-clinical populations^20,21^.

Cognitive changes reflect structural and functional changes in the brain. Healthy age-related changes occur in the thickness of the grey matter mantle (cortical thickness, CT) as well as the widening of the sulci (Sulcal width, SW)^22^, as inferred from structural magnetic resonance imaging (sMRI). SW has recently received increased attention as a robust measurement of cortical morphometry, most notably in older adults^22^ as it appears to be less susceptible to age-related deterioration of sMRI contrast between white and grey matter^23^. A growing number of studies have investigated the brain-associations of cognitive performance of older adults using this technique, with results suggesting that greater sulcal width is associated with poorer cognition and Alzheimer’s disease progression^22, 24, 25^. However, the match between specific online versus in-person domains (working memory, executive functioning, and language) remains unclear, as is the relationship between online cognitive testing and changes in brain structure.

Here we compared the relationship between brain morphology (sulcal width) and cognitive functioning, using both online and in person modalities. We first studied the specific domain-to-domain mapping (working memory, executive functioning, language) between online and in-person testing in a sample (N=140) of healthy adults and then studied the relationship of the latter to cortical morphology as assessed with SW. Furthermore, we sought to disentangle the influence of age, sex, β-amyloid (Aβ) status and genetic risk for Alzheimer’s disease (*APOE*-status) on these associations. We used a partial least square (PLS) multivariate analysis, to reduce the variables to a smaller set of predictors. PLS first extracts a set of latent factors that maximize the covariance between two data sets, here cognition and cortical morphology. These can then be regressed against independent variables such as age, sex and genetic risk.

## METHOD

Data collection and analysis followed approval from the Human Research Ethics committees of QIMR Berghofer (P2193 and P2210), the University of Queensland (2016/HE001261), The University of Newcastle (H-2020-0439) and the National Statement on Ethical Conduct in Human Research. Written informed consent was obtained from all participants following local institutional ethics approval.

### Participants

The 141 participants (75% female, aged 46-71 years, mean age=60; years of education: 13.1 (6.5), NART-IQ mean=110 (SD=9), with 50% or more completed cognitive tasks were drawn from the 159 participants that had attempted online and in-person testing, within the Prospective Imaging Study of Ageing (PISA) cohort-a mid-life cohort, genetically enriched for risk of AD^1^, with data acquired at QIMR Berghofer and Herston Imaging Research Facility (HIRF) in Brisbane, QLD, Australia. All participants have had a structural (T1-weighted) MRI scan and at least 50% of the cognitive scores (online and in-person) available (see Table 1 for a demographic overview).

**Table 1:** Participant demographics for PISA participants who completed both the online and in person cognitive assessments.

| **Variable** |  | **Number of participants** | **Percent** |
| --- | --- | --- | --- |
| **Sex** | Female | 107 | 76% |
|  | Male | 34 | 24% |
| **Education** | <12 years | 38 | 27% |
|  | ≥12 years | 103 | 73% |
| **Amyloid Burden** | Positive | 12 | 9% |
|  | Negative | 122 | 87% |
|  | Missing | 7 | 5% |
| **APOE ε4** | Positive | 66 | 47% |
|  | Negative | 38 | 38% |
|  | Missing | 21 | 15% |

### Neuropsychological assessment

#### Online

Participant recruitment has been fully described elsewhere^1^. In brief, PISA online participants had previously volunteered for genetic studies on risk factors or biomarkers for physical and psychiatric traits. They were re-contacted, consented, and recruited into the PISA study then completed an online survey, subsequently they were invited to undertake online cognitive testing by email approach containing an individualised link. The Creyos battery consists of 12 self-administered tasks across memory, executive function, language, and visuo-spatial domains (listed in supplementary table 1 and fully described at https://creyos.com/). Completion of the full battery takes on average 30 minutes.

#### In-person

A subset of participants who had completed the online phase of the PISA study were invited to complete the in person phase. These were eligible individuals, who were designated at high or low genetic risk of Alzheimer’s disease. The comprehensive face to face cognitive battery was administered assessing cognitive domains of executive functioning, memory, language, and visuo-spatial functioning. All neuropsychological tests listed in Lupton et al.^1^ were administered by trained clinical neuropsychologists. Table 2 lists the tests that were included in this analysis. Completion of the full battery took on average two hours to complete.

**Table 2:** In Person cognitive battery

| Domain | Task |
| --- | --- |
| **Memory** | Rey Auditory Verbal Learning Test - Immediate and Delayed<sup>26,27</sup> |
|  | Topographical Recognition Memory Test<sup>28</sup> |
| **Executive Functions** | Stroop Test (Victoria version)<sup>29</sup> |
|  | Word fluency (FAS)<sup>30</sup> |
|  | Digit Span F/B (Wechsler Adult Intelligence Scale - Fourth edition - WAIS-IV)<sup>31</sup> |
|  | Hayling Sentence Completion Test<sup>32</sup> |
|  | Test of Everyday Attention: Telephone Search; Dual Task<sup>33</sup> |
| **Language** | Graded Naming Test<sup>34</sup> |
|  | National Adult Reading Test<sup>35</sup> |
|  | Spontaneous speech - complex scene description<sup>36</sup> |
|  | Category fluency - Animals<sup>30</sup> |
| **Visuo-spatial** | VOSP-cube<sup>37</sup> |

For some variables, scores were inverted so that a high score always signifies better performance (e.g., task accuracy) and a lower score indicates poorer performance (e.g. error rate, reaction time).

### MRI

As part of an extensive imaging protocol, T1-weighted 3D-MPRAGE structural Magnetic Resonance Imaging (sMRI) data were acquired (TE/TR=2.26 ms/2.3 s, TI=0.9 s, FA=8°, 1 mm isotropic resolution, matrix 256 × 240 × 192, BW=200 Hz/Px, 2x GRAPPA acceleration) at 3T on a Biograph mMR hybrid scanner (Siemens Healthineers, Erlangen, Germany). Other MRI modalities including functional, diffusion and spectroscopy sequences were acquired but are not analysed in the present study^1^.

### APOE *ε4*

*APOE* genotype (***ε***4 allele carriers vs non-carriers) was determined from blood-extracted DNA using TaqMan SNP genotyping assays on an ABI Prism 7900HT and analysed using SDS software (Applied Biosystems). **APOE *ε4*** carriers were coded as positive across homozygeous and heterozygous carriers.

### Aβ quantification

Positron emission tomography (PET) scans were performed on a Biograph mMR hybrid scanner (Siemens Healthineers, Erlangen, Germany) with a Fluorine-18 florbetaben ([^18^F]FBB) diagnostic radiotracer with highly selective binding for Aβ in neural tissue. The centiloid cutoff for Aβ positivity was 20 CL. Full details of acquisition and processing pipelines are provided in Lupton et al^1^.

### Data processing and modelling

#### Sulcal Width (SW)

The Morphologist pipeline of the BrainVISA toolbox^38^ was used to extract local measures of brain anatomy from the T1-w MRI. This pipeline identifies 127 cortical sulci, 63 in the right hemisphere and 64 in the left hemisphere. Cortical thickness (CT) around each sulcus and the sulcal width (SW) were extracted; these have both shown promise for the early detection of AD^22,39^. Following Dauphinot et al.^39^, right and left hemisphere measurements were averaged when the same two sulci exist in each hemisphere, resulting in 64 unique measurements (see Supp. Fig. 1 for abbreviations and full labels). The pipeline was applied in a docker image as described in https://github.com/LeonieBorne/morpho-deepsulci-docker.

#### Partial Least Square (PLS)

Partial least squares (PLS) was used to study co-variation between the two cognitive assays (online and in person) and between cognitive and brain changes across mid- and older adulthood. PLS is a multivariate method that identifies modes of common variation between two data sets and ranks these according to their explained covariance. The resulting projections help identify the most important factors, often referred to as latent variables, that link the two sets of data together, to improve understanding of the relationship between them. The Canonical Partial Least Square (PLS) approach^40^, implemented in the Python library scikit-learn^41^, was used. Two datasets were given as inputs: In the first PLS the first consisted of online and the second of in person testing scores and in the second PLS the first input consisted of sulcal anatomy measures (SW of each sulcus) and the second comprised the cognitive test batteries. This method iteratively calculates pairs of latent variables (modes): the first mode corresponds to the pair explaining the most covariance, and so on for ensuing modes. These latent variables enable both online and in person and brain and cognitive loadings respectively (or back-projections), which weight each individual SW or cognitive test according to their contribution to that mode. Higher scores of these loadings correspond to better task performance and wider sulci, respectively.

PLS models including SW were trained separately for the online and the in-person cognitive tests. Effects of age, sex, *APOE*, **Aβ** were analysed post-hoc using appropriate linear statistics (see below). For all analyses, missing values were replaced by the average score across all participants. Sulci width features and neuropsychological measures were excluded if these were missing in more than 50% of participants. Likewise, participants were excluded if they were missing more than 50% of either cognitive measures or sulci width measures. In total, 3 sulci measures were excluded (F.C.L.r.sc.ant., S.GSM., S.intraCing). No participants were excluded. All measures were z-scored by subtracting the mean of these participants and scaling to unit variance before applying the PLS.

The corresponding code is available at https://github.com/LeonieBorne/brain-cognition-pisa.

### Statistics

#### Permutation tests

Permutation tests were used to identify the robustness of the rank ordered PLS modes^42^. These tests consist of randomly shuffling subject labels in one of the data domains (in this case, the cognitive measures dataset) to disrupt the empirical association with the other domain (sMRI). Then PLS is performed on these shuffled data and the covariance is measured between each pair of latent variables. This test is repeated 1000 times. If the covariance of an empirical mode is greater than 95% of those obtained from the first of these shuffled modes, then that mode is considered robust. As in Smith et al.^43^; we compared scores to the first mode of the permutation tests because this extracts the highest explained variance in a null sample and can thus be viewed as the strictest measure of the null hypothesis^44^.

#### Bootstrapping

Bootstrapping was used to identify which individual measures within a mode had a significant impact on the PLS latent variables^45^. This approach consists of creating a surrogate dataset of the same size as the original data by randomly selecting and removing participants, with replacement. This tests how robust the loadings are to particularities of the original dataset. PLS is then performed on the bootstrapped data and the loadings between each initial measure and the corresponding latent variable are calculated. This test is repeated 1000 times. If the 2.5 and 97.5 percentiles of the loadings obtained have the same sign, the measure (a specific sulcus or cognitive measure) is considered to have a statistically significant impact on the calculation of the latent variable.

#### Statistical analyses

The impact of age was assessed using the Wald Test with the t-distribution as the test statistic. To test whether any age-related effect differs between subgroups (diagnosis, sex, amyloid or *APOE* status), the analysis of covariance (ANCOVA) was used, to test the interaction effect. The effect of sex (male, female) was evaluated using an ANCOVA, controlling for age. The strength of association between in person cognitive testing and sulcal width versus online cognitive testing and sulcal width was tested with Steiger’s z test. The PISA sample was enriched for high genetic risk of AD, including participants who were *APOE* ε4 positive, as well as those in the highest decile of risk for AD as defined by a polygenic risk score (PRS), which was calculated by combining common AD genetic risk variants with *APOE* ε4 omitted (as described in Lupton et al.^1^). To control for any selection bias caused by *APOE* ε4 negative participants being enriched for other AD genetic risk variants, the ANCOVA was also controlled for the AD PRS used in the participant selection.

## RESULTS

### Association between online performance and in-person performance

Across all tests, performance in online cognitive testing strongly and significantly covaried with performance in detailed in-person assessment (cov=2.67; z-cov=12.33; r=0.60; r^2^=0.37; p<0.001; Figure 1). Analyzing different cognitive domains of the face-to face assessment separately (i.e., executive, memory, language and visuo-spatial), revealed that the variance explained for executive tests of the face-to face battery was strongest (cov=1.81; z-cov=11.57; r=0.57; r^2^=0.32; p<0.001), followed by language (cov=1.42; z-cov=7.09; r=0.51; r^2^ =0.26; p<0.001), memory (cov=1.45; z-cov=6.44; r=0.44; r^2^ =0.19; p<0.001), then visuo-spatial (cov=0.44; z-cov=2.60; r=0.26; r^2^=0.07; p=0.013). The average performance on the in-person and online tasks can be found in the supplementary tables 2 and 3.

**Fig. 1:**
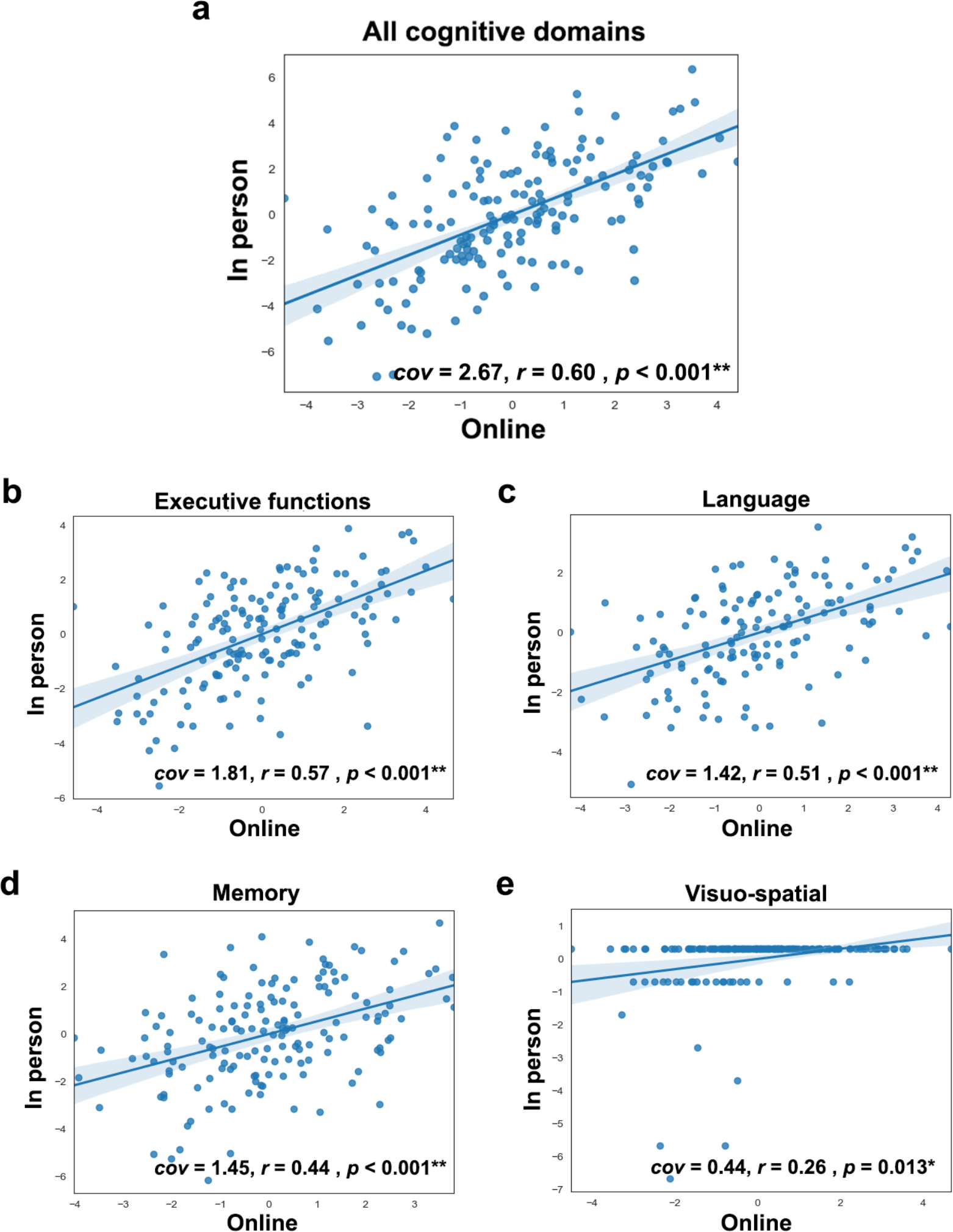
Projections (latent variables) explaining the relationship between online and onsite tests. **a**, Projection (latent variable) of the common variation of all online tests onto all in-person tests. **b**, Projection of online tests of executive function onto in-person tests of executive functions. **c**, Projection of online tests of language onto in-person tests of language. **d**, Projection of online memory tests onto in-person memory tests. **e**, Projection of online tests of visuo-spatial abilities onto in-person tests of visuo-spatial abilities; the shaded area represents the 95% confidence interval.

### Associations between cognition assessments and cortical morphology

The application of partial least square (PLS) yielded a single robust mode for covariation between both the online and in-person assessments, although the nature of the loadings differed somewhat (Fig. 2). The cognitive projection loaded most strongly onto memory and executive functions for the in-person assessment (1st mode, p=0.011, cov=3.55, z-cov=3.00, R2=0.18, z-R^2^=0.95; 2nd mode, p>0.99), and onto executive function for the online battery (1st mode, p<0.001, cov=2.76, z-cov=4.71, R^2^=0.14, z-R^2^=1.15; 2nd mode, p=0.99).

**Figure 2:**
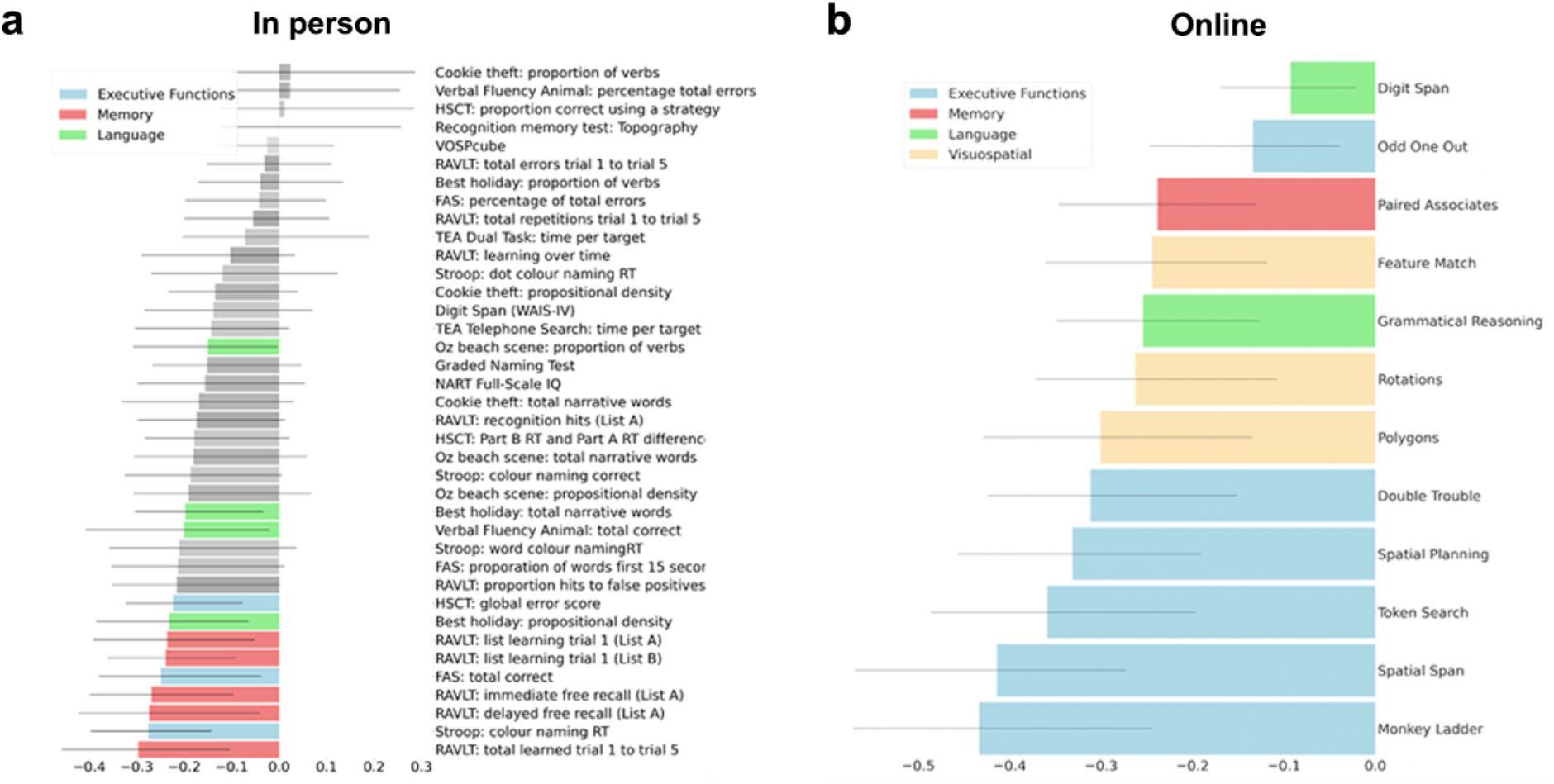
Loadings of the individual cognitive tests of the in person (left) and online (right) battery onto the latent variable of the PLS. **a**, Cognitive test loadings for partial least square (PLS) applied to the in-person assessment, and **b**, to the online assessment. The variables are shown in order of how strongly they load onto the latent variable, with the strongest at the bottom. Tests with non-robust associations are shown in grey.

Brain loadings of cognition-related sulcal width showed a regional pattern that was significantly correlated between the online and in person cognitive appraisals (r=0.996; see figure S1 in supplementary material). Greater SW in these projections covaried with poorer performance in the corresponding cognitive assessments. Brain projections from both cognitive administration modalities (in-person and online) loaded most strongly across the occipital lobe, the anterior and posterior inferior temporal sulcus, the posterior lateral fissure, superior, inferior and internal frontal sulcus, intraparietal sulcus, sub-parietal sulcus, and parieto-occipital fissure.

There was no significant difference in the variance explained in sulci width for the online cognitive assay (r^2^=0.15) compared to the in-person testing (r^2^=0.18; Steiger-z=0.48, p=0.63). Brain-behaviour z-transformed covariance was likewise comparable across the two administration types (Fig. 3).

**Figure 3:**
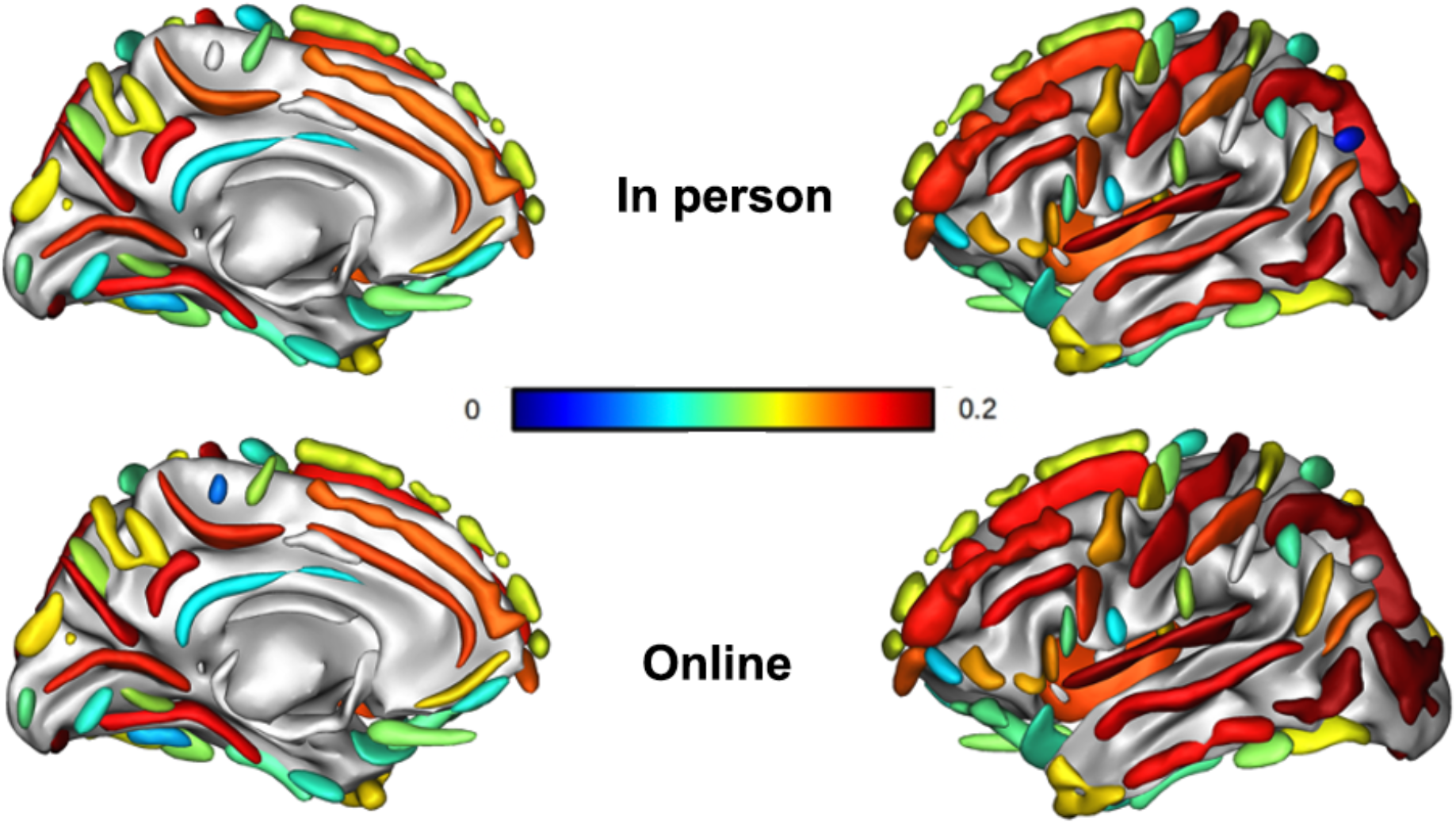
Mean loading of the in person and online latent variable onto the 127 sulci averaged across left and right hemisphere according to BrainVISA toolbox^38^. Mean loading of all reliable sulci in the in-person (top) and online administration (bottom), with the strongest positive covariation of the latent variables of the respective cognitive assays onto the sulcal width latent variable in red and the weakest association in blue.

There was a significant effect of sex on the PLS projections for both the in-person (Fig. 4) and online (Fig. 5) assessments, with poorer performance on the cognitive test (F (1, 140) = 14.89, p<0.001, Fig. 5a) and larger sulcal width for men (F (1,140) = 23.39, p<0.001, Fig. 5d) for the in-person assessment and larger sulcal width for men (F (1, 140) = 21.18, p<0.001, Fig. 4d) in the online assessment. Cognitive performance, both online (F (1,140) = 4.91, p=0.03, Fig. 4b), and in-person (F (1,140) = 8.84, p=0.004, Fig. 5b) was lower for older β-amyloid positive participants. There were no significant age-related differences in sulcal width for either in-person or online assessments and there were no effects of *APOE* ***ε4*** status on cognition or sulcal width, for either the in-person or the online assessment.

**Figure 4:**
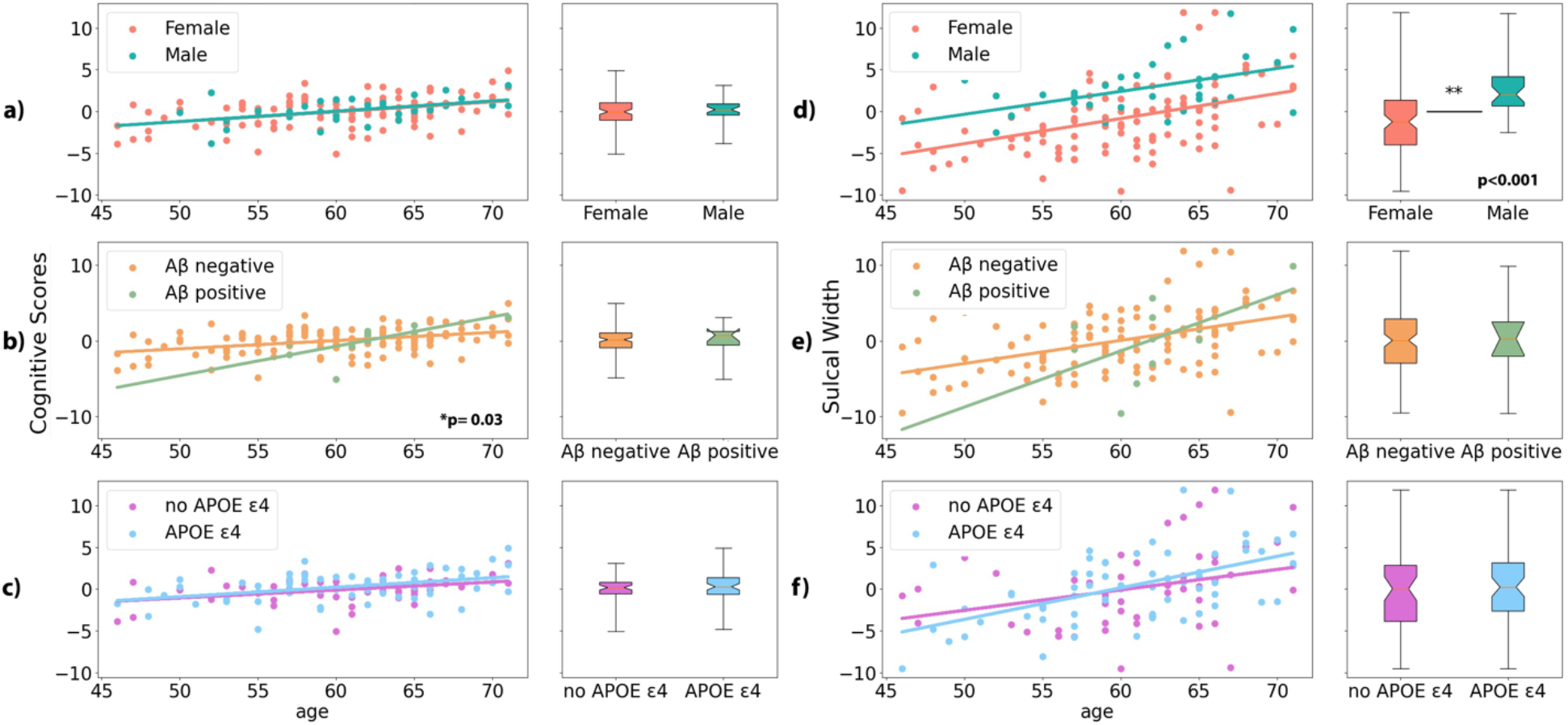
Effects of sex, β-amyloid burden, and *APOE* status on PLS loadings of latent variables of online cognition (left) and sulcal width (right column) Effects of sex (a), amyloid positivity (Aβ) (b), and *APOE* ε*4* status (c) on the cognitive projections. Effects of sex (d), amyloid positivity (Aβ) (e) and *APOE* ε*4* genetic risk (f) on the sulcal width projections in the online condition. Higher scores of these loadings correspond to worse task performance and wider sulci, respectively, significant regressions are indicated by asterisk and p-value.

**Figure 5:**
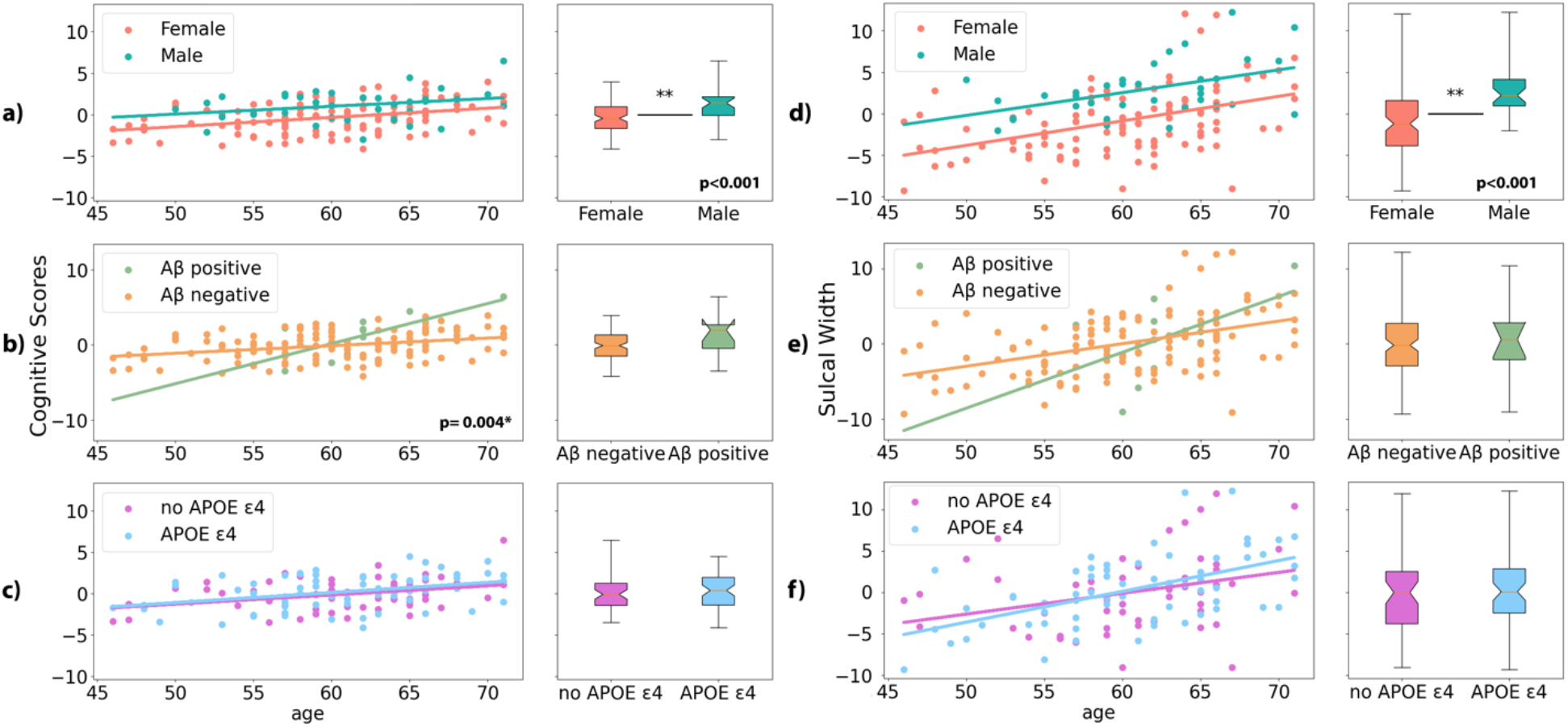
Effects of sex, β-amyloid burden, and *APOE ε4* status on PLS loadings of latent variables of in person cognition (left) and sulcal width (right column). Box and whiskers show mean and inter-quartile range of effects across ages. Effects of sex (a), amyloid positivity (Aβ) (b), and *APOE* ***ε****4* status (c) on the cognitive projections. A higher score on the cognitive loading variable indicates poorer cognition. Effects of sex (d), amyloid positivity (Aβ) (e) and *APOE4* genetic risk (f) on the sulcal width projections in the in-person condition. Higher scores of these loadings correspond to worse task performance and wider sulci, respectively.

## DISCUSSION

With an aging population, and recent advances in treatment options in the early stages of neurodegeneration the demand for early identification is rising. More accessible, digital cognitive testing can assist to fulfill this demand. However, such tests need to have comparable performance to traditional in person tests, and similar sensitivity to the presence and nature of underlying neurobiological differences. Here we demonstrated that relatively brief online cognitive tests strongly co-vary with extensive in-person assessment and relate to similar underlying cortical morphology, with executive and memory domains showing the strongest loadings. A very strong correlation was observed between the sulcal width projections of online and in-person cognitive assays (r=0.996). The variance explained was similarly strong for the online testing (r=0.39) as for the in person testing (r=0.42). For in-person assessment, the cognitive projection loaded strongly onto memory and executive functions, whereas for online the cognitive projection loaded mostly onto executive function. This is likely due to fewer memory-based tests being included in the Creyos platform compared to in-person testing. The strong brain-cognition covariance seen for executive function shows the importance of considering executive function as well as memory when investigating brain neurodegeneration in mid-life aging. In sum, the current analyses suggest adequate sensitivity of online cognitive tests for studying the age-related neurobiology of cognition.

Grouping individuals by demographic or biological variables also shows convergence between online and in person testing. Sex differences were identified on SW projections with lower cognitive test performance and larger sulcal width for men. In previous studies, women have been shown to have higher scores on tests of memory and processing speed compared to men at middle age^46^, in addition to smaller sulcal width which remains consistent across the age spectrum^47^. Our study shows this effect is equally evident in both online and in-person testing. Similarly, the aggregation of Aβ was associated with steeper cognitive age-associated differences in both online and in-person assessments, suggesting that in this healthy community-residing cohort, the impact of early Aβ accumulation is evident in an economical online cognitive assay.

A potential limitation to the study is that the PISA cohort is enriched for those at the extremes of genetic risk for Alzheimer’s disease (selected based on *APOE* genotype and polygenic risk scores). This selection bias does not affect the comparison of the online versus in-person cognitive testing platforms but may predetermine the projections towards prodromal Alzheimer’s disease related impairment, rather than impairment associated with normal aging. The aggregation of Aβ was associated with a poorer cognitive performance for older participants for both cognitive testing modalities, suggesting that in a preclinical sample the early stages of Aβ accumulation accelerate cognitive aging potentially before translation into structural brain changes.

Online cognitive testing has several advantages over in-person cognitive testing, including cost saving, automated interpretation, accessibility, and the ability to adjust the difficulty to match the participant’s ability level^21^. We show that an easily assessable online platform that can be accessed from home and completed unsupervised in 30 minutes has near-comparable prediction of age-associated differences in cortical morphology, compared to an extensive two-hour cognitive test battery administered by a trained neuropsychologist in a controlled environment.

Online testing is being increasingly used for large scale epidemiology studies, such as our PISA study where we have collected data for over 2,000 research participants^1^. It also has the potential to be used as an assessment of the effect of an intervention on cognitive outcomes, and for use as screening tool for the inclusion of participants into clinical trials^48,49^. The latest data collection wave for the Alzheimer’s Disease Neuroimaging Initiative (ADNI), aims to use online screening on 20,000 participants before enrichment for further phenotyping^50^. Ours is the first to directly benchmark online cognitive testing against in person testing in its ability to predict brain morphology. In person cognitive assessment offers alternative strengths, particularly for the clinical assessment and disambiguation of different neurodegenerative disorders in their early phase. In person neurocognitive assessment therefore can assist with diagnosis and treatment tailored to individual patients where it can make a crucial contribution in a clinical setting in the reduction of labor costs and participant time investment.

There are some caveats to note in the current study. There was only a weak correlation between online and in-person performance for the visuo-spatial domains (Fig. 1). This is likely due to the limited variation in scores for the VOSP-cube test which is the sole visuo-spatial task used in the in-person assessment. Overall although we show that both modalities can be used successfully to derive an age effect, the onsite and in-person tests represent each cognitive domain to differing extents, which may have implications in screening for dementia related cognitive decline. Future validation work should also include longitudinal data to allow the effect of change over time to also be assessed.

Unsupervised cognitive testing in a home environment has its own set of limitations that should be taken into account. One of these limitations is the potential for incorrect use of tasks due to various factors, including a lack of understanding of instructions, misinterpretation of instructions, or interruptions and distractions during testing. These issues can affect the accuracy and reliability of the test results and accordingly appropriate measures should be put in place to minimize the impact of these limitations on the testing process, ensuring the quality and reliability of the test results. There is also a risk of intentional misuse such as completion by another individual or purposely failing tasks. Monitoring for validity (such as checking that performance lies within the expected bounds for correct and uninterrupted task completion) may mitigate these issues. This requires consideration if such tests were to be employed as screening tools for inclusion in a clinical trial.

Current alternatives to comprehensive cognitive testing include the Mini-Mental Status Exam (MMSE^51^) and Montreal Cognitive Assessment (MoCA^52^), which are used by health care providers as screening tools and to monitor cognitive changes. Online testing forms, such as Creyos, are likely to become a viable alternative to these tests. The increasing sizes of normative datasets will allow the integration of these platforms into the broader health care system for use by non-experts in monitoring cognitive decline, as well as assessing the effects of drugs or surgery on cognition.

## Data Availability

All data produced in the present study are available upon reasonable request to the authors

## ACKNOWLEDGEMENTS

The Prospective Imaging Study of Aging: Genes, Brain and Behaviour (PISA) was funded by a National Health and Medical Research Council (NHMRC) Boosting Dementia Research Initiative - Team Grant [APP1095227]. We would like to acknowledge and thank all the participants who took part in the study and the PISA team members working on recruitment and data collection.

