## Supplemental Material for "Can an online battery match in-person cognitive testing in predicting age-related cortical changes?"

Supplementary Material

Table S1.: Creyos (Previously Cambridge Brain Sciences) online battery
*Descriptions of tests have been amended from Hampshire et al. (2012) Supplemental information.*

| Domain | | Description |
|---|---|---|
| Memory | Paired Associates | Based on a test commonly used to assess memory impairments in aging clinical populations. Sets of boxes are displayed at random locations on grid. The boxes open one after another to reveal an icon, after which they close. The icons are then displayed sequentially in the centre of the screen, and the participant must select box that contained that icon. If the participant remembers all the icon-location pairs correctly, then the next trial will have one more box. If an error is made the next trial has one less box. The test ends after three errors. The participant's score is the maximum number of pairs successfully remembered. |
| Visuo-spatial | Rotations | Measures the ability to spatially manipulate objects in mind. On each trial, two groups of coloured squares (each with N squares) are displayed beside each other. One of the groups is rotated by a multiple of 90 degrees. The groups are either identical (when un-rotated) or differ by the position of just one item, and participants must indicate if the groups match. They have 90 seconds to complete as many trials as possible. A correct response increases the final score by N, and the subsequent trial has groups of N+1 squares. If the response is incorrect, the total score decreases by N, and next trial has groups of N-1 squares. |
| | Polygons | Based on the Interlocking Pentagons task. On each trial, two overlapping wire-framed polygons are displayed on the left side of screen, and participants must indicate whether the shape to the right is identical to one of the two overlapping ones. A correct response increases the total score by the difficulty level, and the subsequent trial will be more difficult (i.e., differences between polygons will be subtler). An incorrect response decreases the total score by the difficulty level, and the next trial will be slightly easier. |
| | Feature Match | Based on classic feature search tasks used to measure attentional processing. In each trial, two groups of items (each with N items) are displayed beside each other. The groups are either identical in their contents (and item positions), or differ by just one item. Participants have 90 seconds to complete as many trials as possible, indicating whether the groups match. A correct response increases the final score by N, and the subsequent trial has groups of N+1 items. If the response is incorrect, the total |

| | | |
|---|---|---|
| | | score decreases by N, and next trial has groups of N-1 items. |
| Language | Grammatical Reasoning | Based on Alan Baddeley's three minute grammatical reasoning test. On each trial, a written statement regarding two shapes is displayed on the screen, and the participant must indicate whether it correctly describes the shapes pictured below. The participant has 90 seconds to complete as many trials as possible. A correct response increases the total score by one point, and an incorrect response decreases the score by one point. |
| | Digit Span | Based on the verbal working memory component of the WAIS-R intelligence test but only the forward version which is based on the phonological loop of working memory. A sequence of digits is displayed, one at a time, in the centre of the screen. Participants must then repeat the sequence of digits by selecting them on the on-screen keyboard. Difficulty is dynamically varied, and the test ends after three mistakes. The resulting score is the length of the longest digit sequence successfully remembered. |
| Executive function | Token Search | Based on a test used to measure strategy during search behaviour. A set of boxes, one of which contains a hidden token, is displayed on a grid. Participants must find the token by clicking the boxes one at a time. Once found, it is hidden within another box. The token will not appear within the same box twice, thus the participant must search the boxes until the token has been found once within each box. An error is made if the participant checks a box that has: 1) already been clicked while trying to find the token, or 2) previously contained the token. If the participant makes an error, a new trial begins with one less box to search. If the token is found once in each box without any errors being made, a new trial begins with one more box to search. The test finishes after three errors. The resulting score is the maximum level completed. |
| | Double Trouble | Variant of the Stroop test. Either the word "RED" or "BLUE" is displayed on the screen in either the colour red or the colour blue. The participant must select the probe word that correctly describes the colour of the target word. Participants have 90 seconds to complete as many trials as possible. A correct response increases the total score by one, and an incorrect response decreases the score by one. |
| | Odd One Out | Based on a sub-set of problems from the Cattell Culture Fair Intelligence Test. Nine groups of coloured shapes are displayed in a grid. The features define each group (colour, shape, # of items) and are related to each other according to a set of rules. Participants must deduce the rules and select the group whose contents do not correspond to those rules. They have 90 seconds to solve as many problems as possible, and the puzzles get progressively more difficult. A correct response |

| | |
|---|---|
| | increases the final score by one, whereas an incorrect response decreases the score by one. |
| Monkey Ladder | Based on a task from the non-human primate literature. Numbered boxes are displayed at random locations within a grid. After a variable interval (number of squares * 900 ms) the numbers disappear leaving only the boxes. Participants must click the boxes in ascending numerical sequence. The test finishes after three errors, and the resulting score is the length of the longest sequence successfully remembered. |
| Spatial Span | Based on the Corsi Block Tapping Task. 16 purple boxes are displayed in a grid. A sequence of randomly selected boxes turn green one at a time (900 ms per green square). Participants must then repeat the sequence by clicking boxes in the same order. Difficulty is varied dynamically: correct responses increase the length of the next sequence by one square, and an incorrect response decreases the sequence length. The test finishes after 3 errors. The score is the length of the longest sequence successfully remembered. |
| Spatial Planning | Based on the Tower of London Task. Numbered beads are positioned on a tree, and the participant must relocate the beads so that they are arranged in ascending numerical order. They have three minutes to solve as many puzzles as possible, which become progressively harder,requiring more moves and more complex planning. Trials are aborted if the participant makes more than twice the number of moves required to solve the problem. A successfully completed puzzle increases the final score by: (2 x minimum number of moves required) minus the number of moves made. |

Table S2.: Descriptives of the Online tasks

| | N | Mean | Std. Deviation |
|---|---|---|---|
| SpatialSpan | 158 | 4.92 | 0.87 |
| Rotations | 155 | 56.67 | 33.07 |
| Polygons | 153 | 28.12 | 18.39 |
| Paired Associates | 154 | 4.34 | 0.96 |
| Digit Span | 155 | 5.95 | 1.42 |
| Feature Match | 157 | 90.20 | 23.51 |
| Spatial Planning | 145 | 11.87 | 6.66 |
| Grammatical Reasoning | 159 | 13.07 | 4.50 |
| Token Search | 157 | 6.42 | 2.17 |
| Double Trouble | 150 | 13.43 | 13.15 |
| Odd One Out | 154 | 10.29 | 3.08 |
| Monkey Ladder | 155 | 6.97 | 1.13 |

Table S3: Descriptives of the in-person tasks

| | N | Mean | Std. Deviation |
|---|---|---|---|
| NART Full Scale IQ | 159 | 109.60 | 8.44 |
| Graded Naming Test | 159 | 21.89 | 3.40 |
| Oz beach scene total narrative words | 139 | 122.15 | 34.91 |
| Oz beach scene propositional density | 139 | 0.20 | 0.02 |
| Oz beach scene proportion of verbs | 139 | 0.40 | 0.07 |
| Best holiday total narrative words | 139 | 144.02 | 29.93 |
| Best holiday propositional density | 139 | 0.19 | 0.02 |
| Best holiday proportion of verbs | 139 | 0.45 | 0.06 |
| Cookie theft total narrative words | 140 | 154.96 | 34.44 |
| Cookie theft propositional density | 140 | 0.20 | 0.03 |
| Cookie theft proportion of verbs | 140 | 0.49 | 0.06 |
| Animal total correct | 159 | 24.60 | 5.66 |
| Animal percentage total errors | 159 | -2.94 | 3.96 |
| Stroop dot colour naming reaction time | 156 | -12.48 | 2.60 |
| Stroop word colour naming reaction time | 157 | -15.73 | 3.49 |
| Stroop colour naming reaction time | 157 | -25.16 | 6.75 |
| Stroop colour naming correct | 157 | 23.24 | 1.49 |
| HSCT RT B-A difference | 158 | -26.16 | 24.74 |
| HSCT global error score | 158 | -5.69 | 4.09 |
| HSCT proportion correct using a strategy | 158 | 0.49 | 0.25 |
| FAS total correct | 159 | 43.11 | 11.99 |
| FAS proportion of words in first 15 seconds | 159 | -0.40 | 0.07 |
| FAS percentage of total errors | 159 | -6.38 | 4.51 |
| Digit Span WAIS-IV | 159 | 19.72 | 4.02 |
| TEA Telephone Search time per target score | 156 | -3.07 | 0.66 |

| | | | |
|---|---|---|---|
| TEA Dual Task time per target score | 153 | -4.38 | 1.68 |
| RAVLT list learning trial 1 List A | 159 | 6.25 | 1.78 |
| RAVLT total learned trial 1 to 5 | 159 | 50.22 | 9.08 |
| RAVLT total errors trial 1 to 5 | 159 | -1.12 | 1.61 |
| M.RAVLT total repetitions trial 1 to 5 | 159 | 3.40 | 3.42 |
| RAVLT learning over time | 159 | 18.99 | 6.42 |
| RAVLT list learning trial 1 List B | 159 | 5.52 | 1.86 |
| RAVLT immediate free recall List A | 159 | 10.25 | 2.80 |
| RAVLT delayed free recall List A | 159 | 10.21 | 2.96 |
| RAVLT recognition memory hits List A | 159 | 13.67 | 1.46 |
| RAVLT proportion hits to false positives | 159 | 0.91 | 0.10 |
| Recognition memory test Topography | 159 | 26.08 | 2.74 |
| VOSP cube | 159 | 9.69 | 1.00 |

Figure S1: Comparison of sulci-loadings online (left) and in-person (right)(English references for French sulci nomenclature is provided in the Brainvisa Sulci Atlas below)

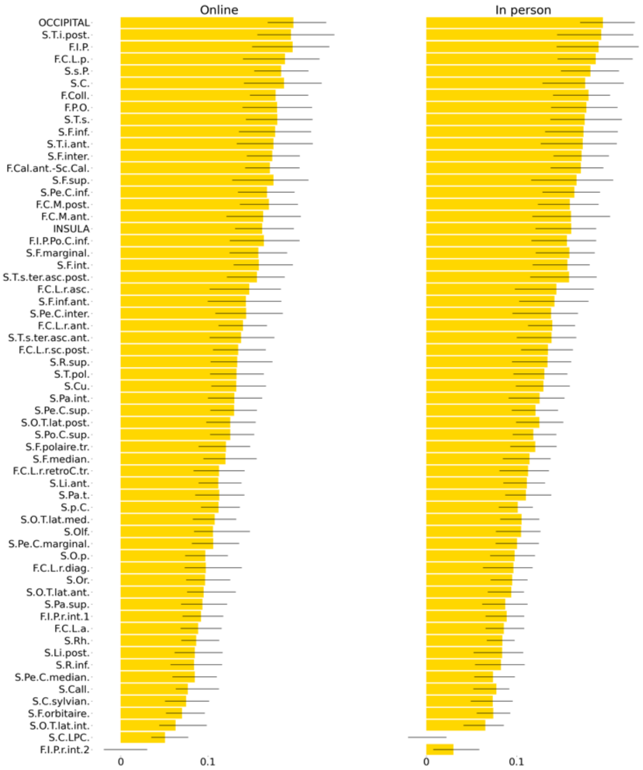


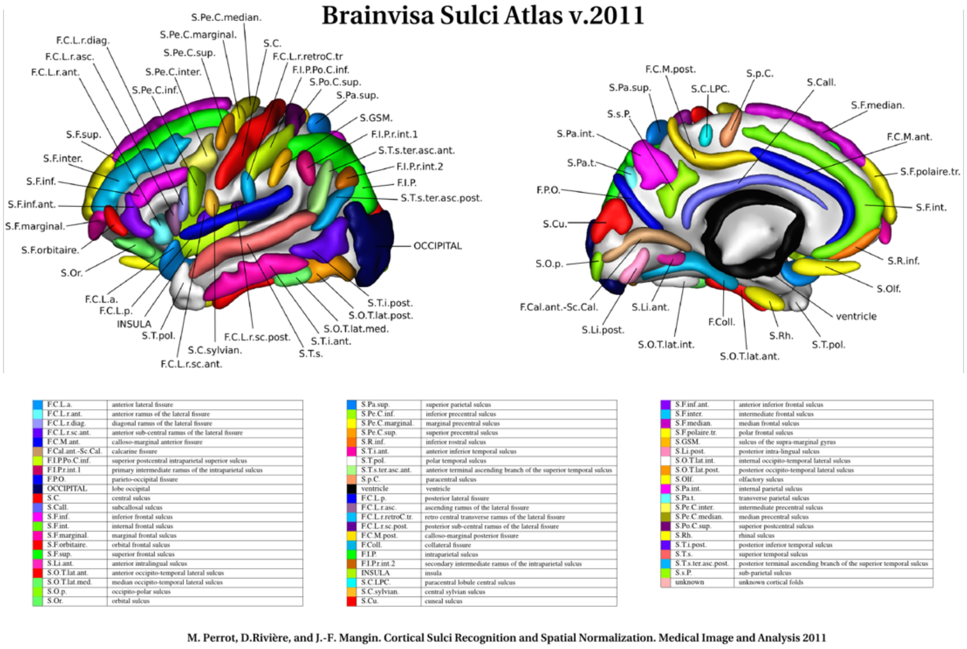
